# Cohort profile: a multicenter European study of acute respiratory infections: the MERMAIDS-ARI 2.0 cohort study

**DOI:** 10.1101/2024.01.16.24301377

**Authors:** Marieke L.A. de Hoog, Cristina Prat-Aymerich, Darren P.R. Troeman, James Lee, Greet Ieven, Peter Horby, Frank Leus, Marc J. M. Bonten, Herman J.A. Goossens, Patricia C.J.L. Bruijning-Verhagen

**Author notes:** **Corresponding Author** Marieke LA de Hoog (E), Julius Centre for Health Sciences and Primary Care, Department of Epidemiology, University Medical Centre Utrecht, Utrecht, The Netherlands, PO Box 85500, 3508 GA Utrecht.

## Abstract

**Purpose:** Multi-centre EuRopean study of MAjor Infectious Disease Syndromes – Acute Respiratory Infections (MERMAIDS-ARI) cohort study is a European prospective cohort study. It was originally launched in 2016 as MERMAIDS-ARI to study acute respiratory infections. When the COVID-19 pandemic hit, the study re-started inclusion under an amended protocol as MERMAIDS-ARI 2.0 focusing on SARS-CoV-2. The objectives of this study were to describe the disease spectrum, clinical features, and outcomes of SARS-CoV-2 infections in hospital care, to determine relevant risk factors and the assess prevalence and within hospital spread compared to other respiratory infections. An important second objective was to establish an extensive biobank supplemented with detailed clinical data to be able to address future virological, immunological, and clinical research questions.

**Methods:** Patients with either 1) ARI presenting to hospital care during the SARS-CoV-2 epidemic (including both COVID-19 and non-COVID-19 patients) or 2) confirmed COVID-19 infection, but with atypical presentation (non-ARI) or with nosocomial acquisition were included. Clinical data and biological samples were collected on enrolment day and repeatedly thereafter until discharge. A subset of COVID-19 patients was also followed up to 12 months post-discharge to study clinical recovery and long-term sequelae.

**Findings to date:** In total 297 patients from 8 countries and 11 sites were recruited between June 2020 and September 2021. We reached high sample completeness for most sample types (>97% for single samples and >86% for serial samples). 294 (99.0%) patients were admitted to hospital with SARS-CoV-2. The population primarily consists of patients (median age 61 years) of Caucasian ethnicity, with the majority being male (62.0%) and having ≥1 comorbidities (67.3%). The median hospital stay was 10 days, and most patients (87.5%) received treatment with systemic corticosteroids and respiratory support (81.4%). A total of 64 (21.7%) patients were admitted to the Intensive Care Unit (ICU), and 6.5% died during hospitalization. Long-term follow-up of 94 patients revealed that most individuals resumed their regular activities and work within three months, with improvements observed in mobility and personal care over time. Instances of severe and extreme pain and anxiety were rarely reported (≤1.1%).

**Conclusion:** This pan-European study offers a valuable resource for conducting in-depth virological, immunological, and pathophysiological investigations pertaining to SARS-CoV-2.

## INTRODUCTION

The COVID-19 pandemic caused a massive influx of patients in hospitals and intensive care units around the world and led to enormous pressures on healthcare systems.

Newly emerging infectious diseases require well-documented patient cohorts rich in biological samples to unravel immunological and pathophysiological disease processes, identify predictive biomarkers and novel targets for therapeutic interventions. In March 2020, the Rapid European COVID-19 Emergency Research response (RECoVER) project involving 10 international partners was selected for funding by the European Union under the Horizon 2020 research framework. RECoVER addressed the most urgent questions related to COVID-19 for patient and public health. One of the RECOVER activities involved the relaunch of the adapted Multi-centre EuRopean study of MAjor Infectious Disease Syndromes – Acute Respiratory Infections (MERMAIDS-ARI). This study originated in 2016 within the Platform for European Preparedness Against (Re-) emerging Epidemics (PREPARE) led by the University of Oxford. The primary aim of MERMAIDS-ARI was to identify host and pathogen related determinants of disease severity of ARI of patients presenting in primary of hospital care. To accommodate specific disease and pathogen characteristics of COVID-19, an adapted version of the MERMAIDS-ARI protocol was developed (MERMAIDS-ARI 2.0) in Spring 2020 with specific focus on SARS-CoV-2 infected and hospitalized patients.

This cohort profile provides a detailed description of the established MERMAIDS-ARI 2.0 cohort and biobank. The cohort and its description can be used to identify suitable patient and sample collections for outstanding clinical, virological and immunological research questions related to COVID-19, for biomarker research and to explore novel targets for therapeutic interventions.

### COHORT DESCRIPTION

The MERMAIDS-ARI 2.0 aimed to investigate the clinical, immunological, and virological characteristics of SARS-CoV-2 in hospital care in European countries. In addition, the study was setup to establish a biobank containing appropriate clinical samples for the study of pathogen replication and excretion within the host, of the host responses to infection and therapy over time (including innate and acquired immune responses), and of host genetic variants associated with disease severity.

#### Site selection

The clinical research infrastructure of COMBACTE network (1) was used for site selection providing access to a large patient population across all age groups and different healthcare environments. An eligibility form was implemented for checking inclusion and exclusion criteria in hospitals that: 1) had a local SARS-COV-2 PCR up and running for clinical care; 2) performed tests on regular basis and 3) kept records of PCRs tests performed including test results and patient identifier. In total 8 European countries (Serbia, Estonia, Greece, Montenegro, Italy, Romania, Spain and the Netherlands) with 11 sites participated in the study (Figure 1). The study was approved by Ethic Committees in each participating country.

**Figure 1:**
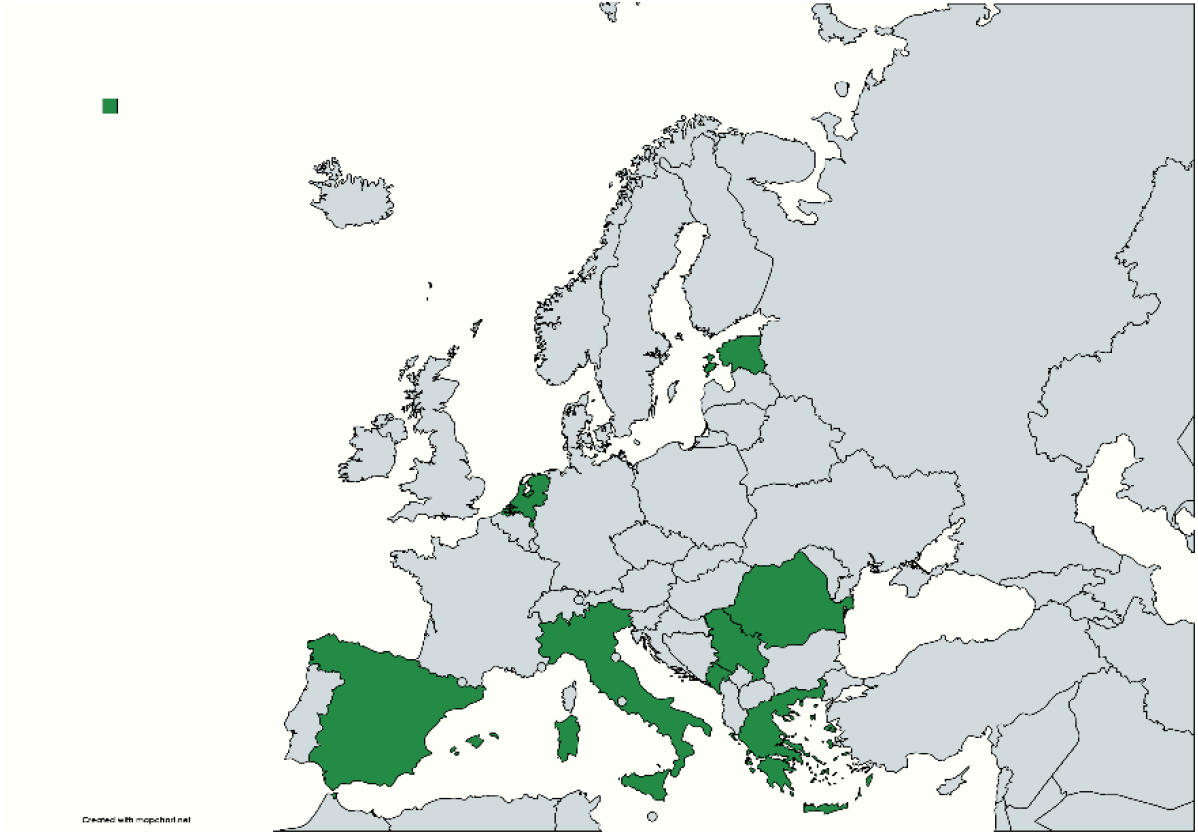
Map of participating countries (www.mapchart.net).

#### Patient enrollment and eligibility

Patient screening and enrollment began in June 2020 and continued until September 2021. Patient eligibility was determined by hospital workers within 48 hours after hospital admission. The patient had to fulfill criteria A or criteria B:

Criteria A: Children or adults admitted to hospital <u>with</u> respiratory symptoms as reason for admission and meeting the following criteria:

1. clinical suspicion of a new episode of ARI with a high probability of SARS-CoV-2 infection (e.g. based on epidemiological linkage or diagnostic procedures);
2. clinical suspicion of ARI as the primary reason for hospital admission, and
3. had onset of the following symptoms within the time window of last 14 days:
  i. Sudden onset of self-reported fever OR temperature of ≥38°C at presentation AND
  ii. At least one respiratory symptom (cough, sore throat, runny or congested nose, dyspnea) AND
  iii. At least one systemic symptom (headache, muscle ache, sweats or chills or tiredness).

Criteria B: Children or adults <u>without</u> respiratory symptoms as reason for admission who had a laboratory confirmed SARS-CoV-2 infection at the time of eligibility check. This allowed for additional identification of (i). severe SARS-CoV-2 cases presented to care but without fever, and (ii). nosocomial cases of SARS-CoV-2.

Microbiological diagnosis was confirmed by molecular testing. If additional tests such as antigen detection were validated and available at the time of inclusion, it was confirmed by a reference method.

Patients who could not or did not provide informed consent, attended primary care but the physician decided to send patient to the hospital for assessment and possible hospital admission or were transferred from another hospital were excluded.

#### Data collection and sampling

At enrollment, patient demographics were collected including sex, ethnicity, educational status, and employment. Detailed clinical information included contact and travel history, medical comorbidities, vaccination status, BMI along with signs and symptoms, vital signs, and laboratory values on admission. Clinical data were subsequently collected at each study visit including signs and symptoms, vital signs, laboratory values, complications, medication, and supportive care received.Serial biological sampling included respiratory, blood and stool specimens at multiple time points. See Table 1 for an overview of the study visit scheme, data collection and biological sampling of MERMAIDS-ARI 2.0.

**Table 1:** Measurements and biological sampling scheme MERMAIDS-ARI 2.0.

|  | Timepoint |  |  |  |  |  |  |  |
| --- | --- | --- | --- | --- | --- | --- | --- | --- |
|  | Hospital admission |  |  |  | Discharge<sup>e</sup> | Post discharge |  |  |
|  | Day 0<sup>a</sup> | Day 2<sup>b</sup> | Day 7<sup>c</sup> | Day 28<sup>d</sup> |  | 3 months | 6 months | 12 months |
| **Questionnaires** |  |  |  |  |  |  |  |  |
| *Baseline characteristics and medical history* | • |  |  |  |  |  |  |  |
| *Employment status* |  |  |  |  |  | • |  |  |
| *Quality of life* |  |  |  |  |  | • | • | • |
| **Clinical data collection<sup>f</sup>** |  |  |  |  |  |  |  |  |
| *Signs and symptoms at admission* | • |  |  |  |  |  |  |  |
| *Vital signs* | • | • | • | • | • |  |  |  |
| *Laboratory values* | • | • | • | • | • |  |  |  |
| *Medication and supportive care* | • | • | • | • | • |  |  |  |
| *Outcome (survival)* |  |  |  |  | • |  |  | • |
| **Biological sampling** |  |  |  |  |  |  |  |  |
| *Blood (EDTA)* | • | • | • |  | • |  |  |  |
| *Serum (SST)* | • |  |  | • |  |  | • | • |
| *NP swab in UTM* | • |  | • | • | • |  |  |  |
| *NP RNA swab* | • |  | • | • | • |  |  |  |
| *Blood RNA (Tempus)* | • | • | • | • | • |  |  |  |
| *Stool sample* |  | • | • | • | • |  |  |  |
| *Blood CPT<sup>g</sup>* |  | • | • | • | • |  |  | • |
| **Pulmonary function<sup>h</sup>** |  |  |  |  |  | • | • |  |

- <sup>a</sup> Day of enrollment - <sup>b</sup> 40 to 56 hours after enrollment. If a participant is discharged on or before day 2; only the discharge samples are collected - <sup>c</sup> If discharged before on or before day 7; discharge samples are collected. - <sup>d</sup> -3 to +15 days if discharged - <sup>e</sup> If discharged within 2 days of sampling for day 7 or day 28; the discharge sample is cancelled. - <sup>f</sup> Information extracted from electronic patient file during hospital stay and at discharge on ICU stay, respiratory and circulatory support, medication and other treatment interventions. - <sup>g</sup> Optional (Tier 2). - <sup>h</sup> including spirometry parameters (TLC, RV, FEV1, FVC, FEV1/FVC ratio) and diffusion capacity (DLCO) according to the guidelines.<sup>2,3</sup>

For all patients, combined nose and throat swab or sample-nasopharyngeal swab were collected at enrollment, day 7, day 28 and at discharge. Stool samples were collected on day 2, 7, 28 and discharge. Blood samples included serum, EDTA and Tempus RNA tubes at enrollment, day 2, day 7, day 28 and at discharge for serology, hematology, blood chemistry as well as host transcriptome studies. Sites could opt-in for collection of additional blood cell-preparation tubes to study adaptive (cellular) immune responses (Tier 2).

Patients were followed until discharge and on day 28 for mortality. In addition, in a subset of sites COVID-19 patients were invited to participate in a post-discharge follow-up with visits at 3-, 6- and 12-months including questionnaires, blood sampling and pulmonary function tests (2, 3) to assess long-term outcomes.

All information was collected in an online eCRF by using the Research Online research data platform. The MERMAIDS-ARI 2.0 data collection was aligned with the ISARIC-CCP data model to improve interoperability and joined data analyses across multiple cohorts.(4) The CRF forms used in MERMAIDS-ARI 2.0 are attached in the supplement.

#### Laboratory testing

All samples were shipped to the central laboratory at University of Antwerp for storage or further processing. SARS-CoV-2 analyses was performed on all sequential NP swabs. Baseline NP swabs were further analyzed by PCR using a respiratory panel that included the following viruses: influenza A, B, A-H1 and A-H3, human coronaviruses NL63, 229E, OC43 and HKU1, Middle East respiratory syndrome coronavirus, severe acute respiratory syndrome coronavirus 1 and 2, parainfluenza viruses 1, 2, 3 and 4, human metapneumovirus, rhinovirus, respiratory syncytial virus A and B, adenovirus, enterovirus, enterovirus D68, parechovirus, and bocavirus. And included the following bacteria: Bordetella pertussis, Chlamydophila pneumoniae, Legionella pneumophila, Mycoplasma pneumoniae, Staphylocococcus aureus, Haemophilus influenzae and Streptococcus pneumoniae. Details of the methods have been described elsewhere.(5)

Peripheral Blood Mononuclear Cells (PBMCs) were isolated from the blood collected at day 2, 7, 28 and discharge in Cell Preparation Tubes (CPT) (Becton Dickinson Diagnostics, Franklin Lakes, NJ, USA) by density gradient centrifugation. Afterwards, they were washed twice with RPMI (Biowest, Nuaillé, France) supplemented with 10% fetal bovine serum (FBS; Biowest, Nuaillé, France) and finally counted using trypan blue in an inverted microscope. For cryopreservation, cells were suspended in 10% DMSO FBS (Sigma-Aldrich, Saint Louis, MO, USA), transferred into cryovials, and then stored at ™80 °C in a cold Nalgene Mr. Frosty Cryo 1 °C Freezing Container (ThermoFisher, Waltham, MA, USA). Cells were then transferred to liquid nitrogen within a week.

Left-over baseline sample NP, PBMC’s, as well as all other samples were biobanked at -80^*^Celsius.

### COHORT CHARACTERISTICS

A total of 297 patients were enrolled in MERMAIDS-ARI 2.0 between June 2020 and September 2021. Of the 297 patients, 294 (99.0%) were admitted to hospital with SARS-CoV-2 and for 237 patients this was confirmed in the baseline NP swab upon study inclusion. Figure 2 provides an overview of monthly patient enrolment over time. At least one biological sample was obtained from 290 patients (97.6%). Per sample type, completeness varied between 56.9% for at least one stool sample to 97.6% for at least one blood sample and NP swab. Completeness was highest for baseline samples (> 95.6%, except for stool). Serial samples were available for 35.4% (stool) to 92.6% (blood) of sample types (Table 2).

**Table 2:**
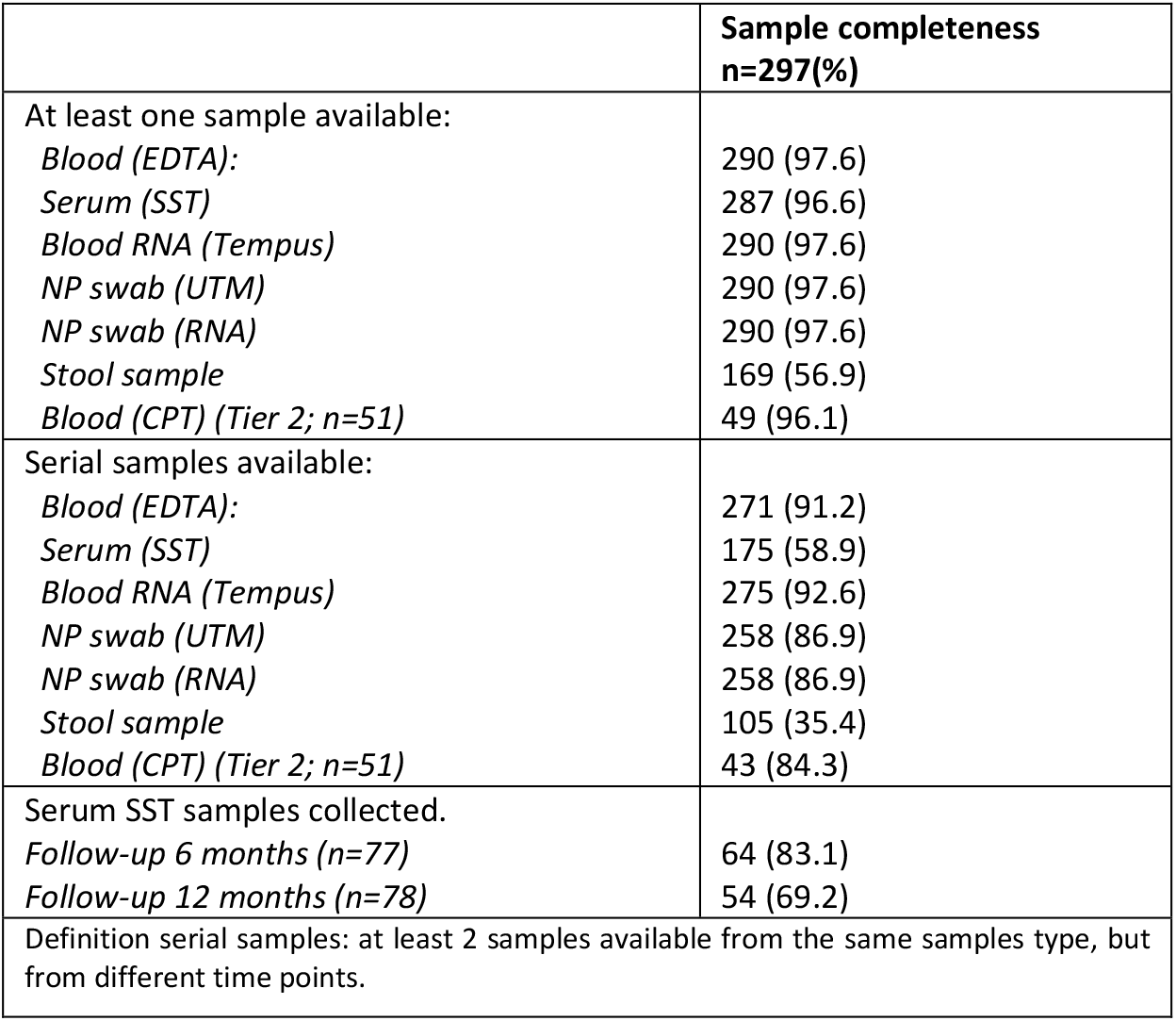
Sample completeness (hospital) sites of MERMAIDS-ARI 2.0.

|  | Sample completeness<br>n=297(%) |
| --- | --- |
| At least one sample available: |  |
| *Blood (EDTA):* | 290 (97.6) |
| *Serum (SST)* | 287 (96.6) |
| *Blood RNA (Tempus)* | 290 (97.6) |
| *NP swab (UTM)* | 290 (97.6) |
| *NP swab (RNA)* | 290 (97.6) |
| *Stool sample* | 169 (56.9) |
| *Blood (CPT) (Tier 2; n=51)* | 49 (96.1) |
| Serial samples available: |  |
| *Blood (EDTA):* | 271 (91.2) |
| *Serum (SST)* | 175 (58.9) |
| *Blood RNA (Tempus)* | 275 (92.6) |
| *NP swab (UTM)* | 258 (86.9) |
| *NP swab (RNA)* | 258 (86.9) |
| *Stool sample* | 105 (35.4) |
| *Blood (CPT) (Tier 2; n=51)* | 43 (84.3) |
| Serum SST samples collected. |  |
| *Follow-up 6 months (n=77)* | 64 (83.1) |
| *Follow-up 12 months (n=78)* | 54 (69.2) |
| Definition serial samples: at least 2 samples available from the same samples type, but from different time points. |  |

**Figure 2:**
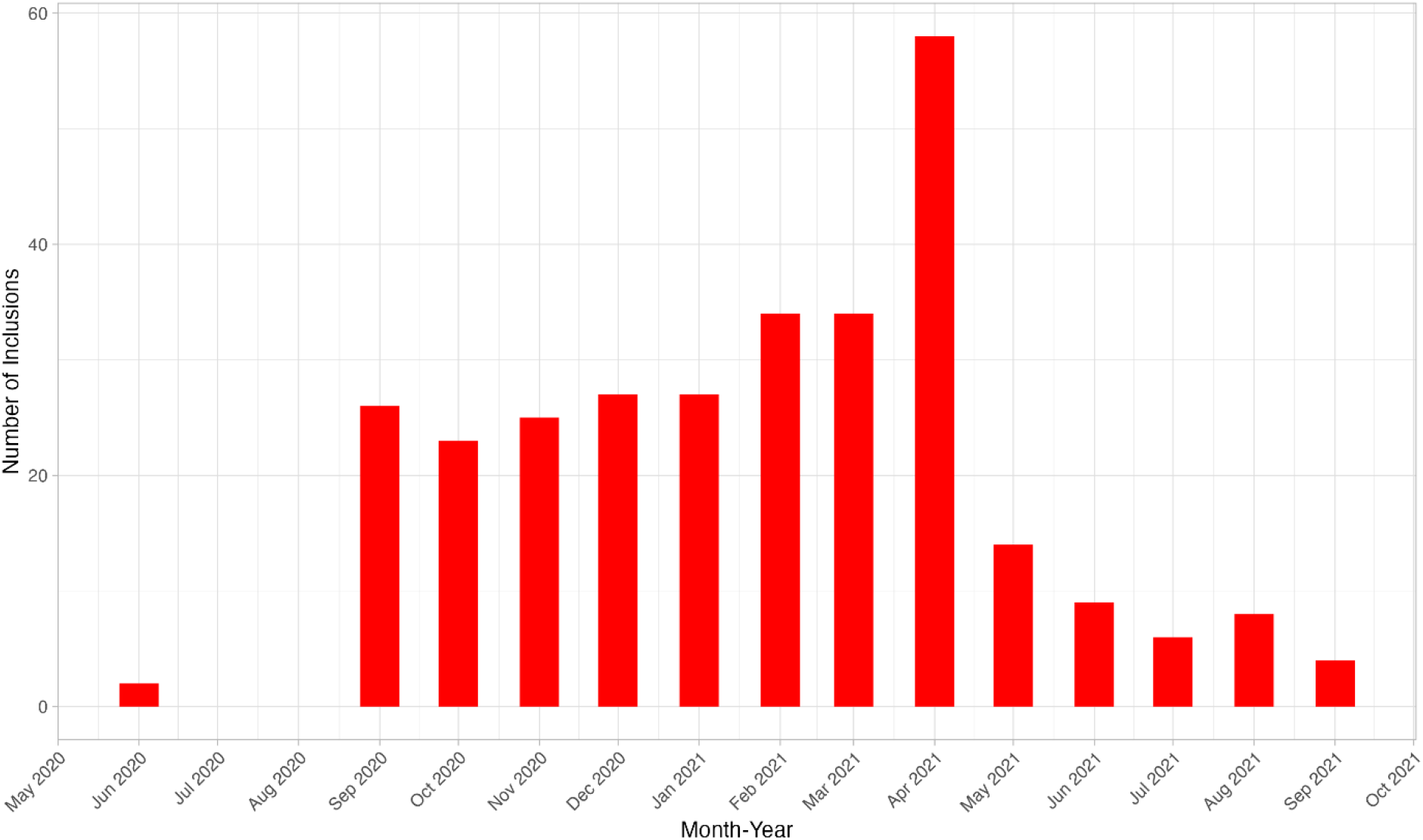
Monthly inclusions in the MERMAIDS-ARI 2.0 study patients.

#### Patient characteristics and medical history

Table 3 shows baseline characteristics of the 297 patients. More males than females were included (62.0% males and 38.0% females). The majority was of white/Caucasian ethnicity (95.6%). The median age of the hospitalized patients was 61 year and one child (5-year-old) was included in the study.

**Table 3:** Patient demographics and medical history by SARS-CoV-2 result and hospital versus GP.

|  | n=297(%) |
| --- | --- |
| Demographics |  |
| Sex |  |
| *Male* | 184 (62.0) |
| *Female* | 113 (38.0) |
| Median age (range) | 61 (5-89) |
| Race/ethnicity |  |
| *White/Caucasian* | 284 (95.6) |
| *Other* | 12 (4.0) |
| Lives in nursing home/long-term healthcare facility |  |
| *Yes* | 4 (1.3) |
| *No* | 293 (98.7) |
| Education* |  |
| *Completed high school or higher* | 214 (72.1) |
| *Completed primary school* | 40 (13.7) |
| *Unknown* | 43 (14.5) |
| Employment status* |  |
| *Employed* | 123 (41.9) |
| *Unemployed* | 26 (8.9) |
| *Retired* | 126 (43.0) |
| *Homemaker/child care/elder care* | 13 (4.4) |
| *Student* | 3 (1.0) |
| *Other* | 1 (0.3) |
| healthcare worker | 15 (5.4) |
| Smoking history |  |
| *Never* | 210 (70.7) |
| *Current or past smoker* | 86 (29.0) |
| Medical history |  |
| BMI |  |
| *Underweight (<18.5)* | 5 (1.7) |
| *Normal weight (18.5-25)* | 68 (23.0) |
| *Overweight (25-30)* | 109 (36.8) |
| *Obese (≥30)* | 114 (38.5) |
| Modified Charlson comorbidity index(8) |  |
| *0* | 97 (32.7) |
| *1* | 88 (29.6) |
| *2* | 68 (22.9) |
| *≥3* | 44 (14.8) |
| Type of comorbidity (% yes) |  |
| *Chronic cardiac disease (including arterial hypertension)* | 163 (34.1) |
| *Chronic pulmonary disease (excluding asthma)* | 137 (28.7) |
| *Asthma (physician diagnosed)* | 24 (8.1) |
| *Cerebrovascular disease\** | 22 (7.4) |
| *Chronic kidney disease* | 22 (7.4) |
| *Peptic ulcer disease\** | 7 (2.4) |
| *Liver disease* | 13 (4.4) |
| *Chronic neurological disorder* | 19 (6.4) |
| *Malignant neoplasm\** | 23 (7.8) |
| *Chronic hematologic disease\** | 5 (1.7) |
| *Connective tissue disorder* | 23 (7.7) |
| *Diabetes (Type 1 or Type 2)* | 15 (5.1) |
| *Dementia* | 2 (0.7) |
| Prior SARS-CoV-2 infection |  |
| *Yes* | 5 (1.8) |
| *No* | 279 (98.2) |
| Vaccination history |  |
| *Pneumococcal (ever)* | 24 (8.1) |
| *Influenza (within last 6 months)* | 50 (16.8) |
| *COVID-19 (at least one dose)* | 139 (46.8) |
| COVID-19 vaccination type* |  |
| *Pfizer - BioNTech* | 60 (43.2) |
| *Moderna* | 23 (16.5) |
| *AstraZeneca* | 6 (4.3) |
| *Sputnik* | 3 (2.2) |
| *Janssen - Johnson&Johnson* | 14 (10.1) |
| *Sinopharm* | 34 (25.2) |
| COVID-19 vaccination* |  |
| *Single dose* | 22 (15.8) |
| *Completed primary series<sup>#</sup>* | 112 (80.6) |
| *Unknown* | 5 (3.6) |
| Not all numbers add up to the total number of patients due to some missing data. The percentage presented are the valid percentages excluding those patients with missing data.<br>*Among those who have received a COVID-19 vaccination. One person received combination of Pfizer and AstraZeneca vaccine.<br><sup>#</sup>Two doses or one dose with Janssen – Johnson&Johnson |  |

Most patients had ≥1 comorbidity (67.3%). The most reported comorbidities across all patients were obesity (38.5%), chronic cardiovascular disease (34.1%) and chronic pulmonary disease (28.7%).

#### Hospitalization, treatment, and survival

The median hospital stay was 10 days (range 2-71 days; Table 4). Most patients (87.5%) were treated with systemic corticosteroids and received respiratory support (81.4%). In total 64 (21.7%) patients were admitted to the ICU and 28 (9.5%) received mechanical ventilation. Any kind of complication occurred in 57.5% of patients (57.5%) and 19 patients (6.5%) died during hospital admission.

**Table 4:** Treatment during hospital stay.

|  | **n=297(%)** |
| --- | --- |
| Median day of hospital stay (range) | 10 (2-71) |
| Admission to ICU |  |
| *Yes* | 64 (21.7) |
| *No* | 231 (78.3) |
| Median day of ICU admission (range) | 10 (0-34) |
| Received COVID-19 treatment |  |
| *Any systemic corticosteroids* | 258 (87.5) |
| *Dexamethasone* | 148 (57.4) |
| *Methylprednisolone* | 100 (38.8) |
| *Immunomodulatory or antibody based COVID-19 treatment* | 48 (16.3) |
| *Antivirals* | 148 (50.2) |
| Respiratory support |  |
| *No* | 55 (18.6) |
| *Supplemental oxygen only* | 156 (52.9) |
| *Non-invasive ventilation (with or without supplemental oxygen)* | 55 (18.6) |
| *Mechanical ventilation* | 28 (9.5) |
| *ECMO* | 1 (0.3) |
| Not all numbers add up to the total number of patients due to some missing data. The percentage presented are the valid percentages excluding those patients with missing data. |  |

**Table 5:** Complications and survival.

|  | n=297(%) |
| --- | --- |
| **Complications** |  |
| *Any complications* | 168 (57.5) |
| *Cardiac complications<sup>a</sup>* | 41 (14.2) |
| *Thromboembolic or coagulation complications<sup>b</sup>* | 49 (26.9) |
| *Other complications<sup>c</sup>* | 135 (47.4) |
| **Survival** |  |
| *Alive at last visit or discharge* | 275 (93.5) |
| *Death* | 19 (6.5) |
| • Not all numbers add up to the total number of patients due to some missing data. The percentage presented are the valid percentages excluding those patients with missing data.<br>• <sup>a</sup> Cardiac complications include: heart failure, endocarditis, arrhythmia, ischemia, and cardiac arrest.<br>• <sup>b</sup> Thromboembolic/coagulation complications include pulmonary embolism, deep venous thrombosis, stroke, and coagulopathy/disseminated intravascular coagulation.<br>• <sup>c</sup> Other complications include: bacterial pneumonia, ARDS, pneumothorax, cryptogenic organizing pneumonia, meningitis, seizure, bacteremia, anemia, rhabdomyolysis, gastrointestinal bleeding, pancreatitis, liver dysfunction, hyperglycemia, hypoglycemia, or other complications. |  |

#### Long term outcomes at 3-, 6- and 12-months

In total 94 COVID-19 patients participated in the post-discharge substudy with follow-up visits at 3 (n=94), 6 (N=77) and 12 months (n=78) after discharge. Table 6 shows the characteristics and the physical and mental recovery of these patients.

**Table 6:** Characteristics and recovery of patients in extended follow-up.

|  | 3 months after<br>discharge<br>n=94(%) | 6 months after discharge<br>n=77(%) | 12 months after<br>discharge<br>n=78(%) |
| --- | --- | --- | --- |
| **Demographics** |  |  |  |
| **Sex** |  |  |  |
| *Male* | 62 (66.0) | 48 (62.3) | 52 (66.7) |
| *Female* | 32 (34.0) | 29 (37.7) | 26 (33.3) |
| Median age (range) | 58 (5-89) | 58 (18-81) | 57 (5-81) |
| **Ethnicity** |  |  |  |
| *White/Caucasian* | 92 (97.9) | 75 (97.4) | 77 (98.7) |
| *Other* | 2 (2.1) | 2 (2.6) | 1 (1.3) |
| **Employment status** |  |  |  |
| *Employed* | 48 (53.9) | - | - |
| *Employed (paid sick leave)* | 3 (3.4) | - | - |
| *Unemployed* | 6 (6.7) | - | - |
| *Retired* | 25 (28.1) | - | - |
| *Student* | 2 (2.2) | - | - |
| *Home maker/ childcare / elder care* | 4 (4.5) | - | - |
| *Unknown* | 1 (1.1) | - | - |
| Return to work and usual activities in first 3 months after discharge |  |  |  |
| Return to usual activities (unemployed; n=37) |  |  |  |
| 1 week | 2 (5.4) | - | - |
| 2 weeks | 9 (24.3) | - | - |
| 4 weeks | 13 (35.1) | - | - |
| 8 weeks | 7 (18.9) | - | - |
| 12 weeks | 1 (2.7) | - | - |
| Activities not resumed yet | 5 (13.5) | - | - |
| Return to work (employed; n=53) |  |  |  |
| 1 week | 3 (5.7) | - | - |
| 2 weeks | 19 (35.8) | - | - |
| 3 weeks | 2 (3.8) | - | - |
| 4 weeks | 15 (28.3) | - | - |
| 8 weeks | 8 (15.1) | - | - |
| 12 weeks | 3 (5.7) | - | - |
| Patient has not returned to work | 3 (5.7) | - | - |
| Employed: had to make significant changes to their work duties or occupation because of their hospital stay (% yes) | 6 (11.3) | - | - |
| Effectiveness on the job (0-100) median (range) | 90 (0-100) | - | - |
| Health/welfare |  |  |  |
| Mobility:<br>Problem with walking around |  |  |  |
| No | 56 (59.6) | 49 (64.5) | 61 (78.2) |
| Slight | 20 (21.3) | 18 (23.7) | 11 (14.1) |
| Moderate | 13 (13.8) | 4 (5.3) | 3 (3.8) |
| Severe | 4 (4.3) | 4 (5.3) | 1 (2.6) |
| Unable to walk | 1 (1.1) | 1 (1.3) | 1 (1.3) |
| Personal care:<br>Problem with washing or dressing |  |  |  |
| No | 88 (88.3) | 71 (94.7) | 72 (93.5) |
| Slight | 9 (9.6) | 2 (2.2) | 2 (2.6) |
| *Moderate* | 1 (1.1) | 1 (1.3) | 1 (1.3) |
| *Severe* | 0 (0.0) | 0 (0.0) | 1 (1.3) |
| *Unable to wash or dress* | 1 (1.1) | 1 (1.3) | 1 (1.3) |
| Usual activities:<br>Problem doing usual activities |  |  |  |
| *No* | 65 (69.9) | 58 (76.3) | 61 (78.2) |
| *Slight* | 18 (19.4) | 11 (14.5) | 12 (15.4) |
| *Moderate* | 8 (8.6) | 6 (7.9) | 3 (3.8) |
| *Severe* | 0 (0.0) | 0 (0.0) | 1 (1.3) |
| *Unable to do my usual activities* | 2 (2.2) | 1 (1.1) | 1 (1.3) |
| Having pain or discomfort |  |  |  |
| *No* | 54 (58.1) | 55 (72.4) | 58 (74.4) |
| *Slight* | 24 (25.8) | 16 (21.1) | 16 (20.5) |
| *Moderate* | 14 (15.1) | 3 (3.9) | 3 (3.8) |
| *Severe* | 0 (0.0) | 1 (1.3) | 0 (0.0) |
| *Extreme* | 1 (1.1) | 1 (1.3) | 1 (1.3) |
| Being anxious or depressed |  |  |  |
| *No* | 65 (69.1) | 58 (76.3) | 60 (76.9) |
| *Slight* | 20 (21.3) | 13 (17.1) | 16 (20.5) |
| *Moderate* | 8 (8.5) | 4 (5.3) | 2 (2.6) |
| *Severe* | 1 (1.1) | 1 (0.0) | 0 (0.0) |
| *Extreme* | 0 (0.0) | 1 (1.3) | 0 (0.0) |
| Median health rating (range 0-100) | 80 (20-100) | 85 (30-100) | 85 (8-100) |
| Pulmonary function |  |  |  |
| Pulmonary function measured (% yes) | 32 (34.0) | 42 (54.5) | - |
| Normal lung function measured?* |  |  |  |
| *Yes* | 22 (68.8) | 34 (81.0) | - |
| *No* | 5 (15.6) | 7 (16.7) | - |
| *Unknown* | 5 (15.6) | 1 (1.3) | - |
| Not all numbers add up to the total number of patients due to some missing data. The percentage presented are the valid percentages excluding those patients with missing data.<br>*Among those whose lung function is measured. |  |  |  |

Three months after discharge, most patients had resumed their usual activities (85.6%; the unemployed) or returned to work (94.3%; the employed). Of the employed patients, 11.3% had to make significant changes to their work duties or occupation because of their COVID-19 illness episode. Most of the patients reported no or only slight problems with mobility (no problem: 59.6% and slight problem: 21.3%), personal care (no problem: 88.3% and slight problem: 9.6%), and usual activities (no problem: 69.9% and slight problem: 19.4%). Pain or discomfort and anxiety or depression were not reported very often (16.2% of the patients reported moderate to extreme pain and discomfort and 9.6% reported moderate to extreme anxiety and depression). All parameters improved over time at 6- and 12 months. Median health rating was 80 (range 20-100) on a scale from 0-100 three months after discharge and improved to 85 at 6- and 12 months after discharge.

## DISCUSSION

This longitudinal, hospital-based cohort study enrolled 297 patients from eight different European countries in the COVID-19 pandemic covering infections with the Wild-type, Alpha, and Delta variant. Strengths of our cohort study include the prospective design with detailed and standardized clinical follow-up and the serial sampling resulting in an extensive biobank and the Pan-European coverage. Jointly, this collection enables in-depths mechanistic studies on host-pathogen interactions, pathways for disease progression and virus evolution and has already contributed to some key insights on these topics.(6, 7) Furthermore, data collection was harmonized to international initiatives improving interoperability and joint analyses of multiple cohorts. Requests for individual level patient data and biobank access can be sent to first author (MLAH) who will seek agreement from the core research team.

Planned further analyses include virus-virus and virus-bacterial interactions and transcriptomic analyses for biomarker discovery.

The study also has some limitations. First, we did not succeed in enrolling pediatric subjects into the study. Main reason for this was the very low admission rate for COVID-19 among children, which later led to a protocol adaptation shifting focus entirely to adult patients. Second, we were unable to obtain serial samples from all enrolled subjects, which will impact the sample size available for some research questions. Third, most patients were enrolled in the first half of the pandemic. About half of patients had received at least one dose of COVID-19 vaccination and a limited number received (repeated) booster doses. Also, the proportion of patients with known prior SARS-CoV-2 infection is low. This may impact the ability to generalize (immunological) findings to the current era of SARS-CoV2 endemicity. Similarly, our cohort represents infections with wild-type and early variants of SARS-CoV-2.

## Conclusions

We describe a prospective pan-European hospital based COVID-19 patient cohort and extensive biobank established during the COVID19 pandemic, which can provide a rich source for further virological, immunological, and pathophysiological studies related to SARS-CoV2.

## Data Availability

All data produced in the present study are available upon reasonable request to the authors

## ACKNOWLEDGEMENTS

We want to express our gratitude to the 11 hospital research teams, their doctors/nurses and practice personnel for implementing the study.

## DISCLOSURE STATEMENT

The authors alone are responsible for the content and writing of the paper. The funder had no role in the design and conduct of the study; collection, management, analysis, and interpretation of the data; preparation, review, or approval of the manuscript; and decision to submit the manuscript for publication.

### Ethical statement

Regulatory approvals were sought in each country. The following Ethics Boards were consulted: Medical Ethics Committee of the: University Medical Centre Utrecht-the Netherlands, Clinical Center of Serbia-Serbia, Clinical Centre of Kragujevac-Serbia, Tartu University Clinic-Estonia, Attikon University General Hospital-Greece, Brescia Hospital-Italy, Clinic Center of Podgorcia-Montenegro, National Institute for Infectious Diseases ‘Prof. Dr. Matei Bals’-Romania, University Hospital Brothers Trias I Pujol-Spain, HU Virgen Macarena-Spain and the Regional Hospital de M Laga-Spain. Dates of approval letters and/or numbers available on request.

